# Distinct systems serology features in children, elderly and COVID patients

**DOI:** 10.1101/2020.05.11.20098459

**Authors:** Kevin J. Selva, Carolien E. van de Sandt, Melissa M. Lemke, Christina Y. Lee, Suzanne K. Shoffner, Brendon Y. Chua, Thi H.O. Nguyen, Louise C. Rowntree, Luca Hensen, Marios Koutsakos, Chinn Yi Wong, David C. Jackson, Katie L. Flanagan, Jane Crowe, Allen C. Cheng, Denise L. Doolan, Fatima Amanat, Florian Krammer, Keith Chappell, Naphak Modhiran, Daniel Watterson, Paul Young, Bruce Wines, P. Mark Hogarth, Robyn Esterbauer, Hannah G. Kelly, Hyon-Xhi Tan, Jennifer A. Juno, Adam K. Wheatley, Stephen J. Kent, Kelly B. Arnold, Katherine Kedzierska, Amy W. Chung

## Abstract

SARS-CoV-2, the pandemic coronavirus that causes COVID-19, has infected millions worldwide, causing unparalleled social and economic disruptions. COVID-19 results in higher pathogenicity and mortality in the elderly compared to children. Examining baseline SARS-CoV-2 cross-reactive coronavirus immunological responses, induced by circulating human coronaviruses, is critical to understand such divergent clinical outcomes. The cross-reactivity of coronavirus antibody responses of healthy children (n=89), adults (n=98), elderly (n=57), and COVID-19 patients (n=19) were analysed by systems serology. While moderate levels of cross-reactive SARS-CoV-2 IgG, IgM, and IgA were detected in healthy individuals, we identified serological signatures associated with SARS-CoV-2 antigen-specific Fcγ receptor binding, which accurately distinguished COVID-19 patients from healthy individuals and suggested that SARS-CoV-2 induces qualitative changes to antibody Fc upon infection, enhancing Fcγ receptor engagement. Vastly different serological signatures were observed between healthy children and elderly, with markedly higher cross-reactive SARS-CoV-2 IgA and IgG observed in elderly, whereas children displayed elevated SARS-CoV-2 IgM, including receptor binding domain-specific IgM with higher avidity. These results suggest that less-experienced humoral immunity associated with higher IgM, as observed in children, may have the potential to induce more potent antibodies upon SARS-CoV-2 infection. These key insights will inform COVID-19 vaccination strategies, improved serological diagnostics and therapeutics.

## Introduction

Since the first reported coronavirus disease 2019 (COVID-19) patient in December 2019^1^, the severe acute respiratory syndrome coronavirus 2 (SARS-CoV-2) has become a global pandemic, infecting millions of individuals worldwide^2^. Though the majority of COVID-19 patients experience mild symptoms, approximately 20% of cases have more severe disease outcomes involving hospitalization or intensive care treatment, especially in those with underlying co-morbidities such as diabetics and cardiovascular disease^3^. Furthermore, COVID-19 related morbidity and mortality is significantly higher in the elderly population and almost absent in school-aged children^4^. A disproportional outcome in disease severity with increasing age is not unique to the SARS-CoV-2 pandemic, and is observed during previous influenza pandemics^5^. Understanding whether baseline pre-existing immunological responses, induced by previous exposure to seasonal coronaviruses, contributes to such differences may provide important insights to such divergent clinical outcomes between children and elderly.

Antibodies (Abs) are a vital component of the immune response with demonstrated importance in the control of most viral pathogens. Apart from virus neutralization, Abs have the capacity to engage Fc Receptors (FcRs) or complement to induce Fc effector functions, including Ab-dependent cellular cytotoxicity (ADCC), Ab-dependent cellular phagocytosis (ADCP), or Ab-dependent complement activation (ADCA)^6^. Fc functions are not limited to neutralizing viral epitopes but utilize any available epitope^6^. A previous SARS-CoV (also called SARS-CoV-1) study associated ADCP with viral clearance^7^, where individuals expressing a higher affinity FcγRIIa-H131 polymorphism, associated with enhanced Fc functions, had better disease outcomes^8^. However Fc functional Abs may also enhance infection or pathology through Ab-dependent enhancement (ADE), previously observed in some SARS-CoV-1 animal vaccine and *in vitro* studies^9,10^. Hence, there is an urgent need to understand the Ab responses elicited against SARS-CoV-2, especially given that current COVID-19 vaccine strategies are focused upon inducing effective neutralizing Ab responses^11^, without inducing pathological damage^12^.

## Results

### Immature humoral immunity in children

In-depth characterization of cross-reactive SARS-CoV-2 Ab responses in healthy children compared to healthy elderly is needed to understand whether pre-existing human coronavirus (hCoV)-mediated Ab immunity potentially contributes to COVID-19 outcome. We designed a cross-reactive CoV multiplex array, including SARS-CoV-2, SARS-CoV-1, MERS-CoV and hCoV (229E, HKU1, NL63) spike (S) and nucleoprotein protein (NP) antigens **(Extended Data Figure 1b)**. CoV-antigen-specific levels of isotypes (IgG, IgA, IgM) and subclasses (IgG1, IgG2, IgG3, IgG4, IgA1, IgA2), along with C1q binding (a predictor of ADCA via the classical pathway) and FcγRIIa, FcγRIIb and FcγRIIIa soluble dimer engagement (mimicking FcγR immune complex formation associated with the induction of a range of Fc effector functions^13^) were assessed from plasma of 89 children, 98 adults, and 57 elderly individuals **(Extended Data Figure 2a and Extended Data Table 1)**, generating a composite dataset of baseline CoV Ab features (14 CoV antigens x 14 detectors = 196 Ab features).

**Figure 1.**
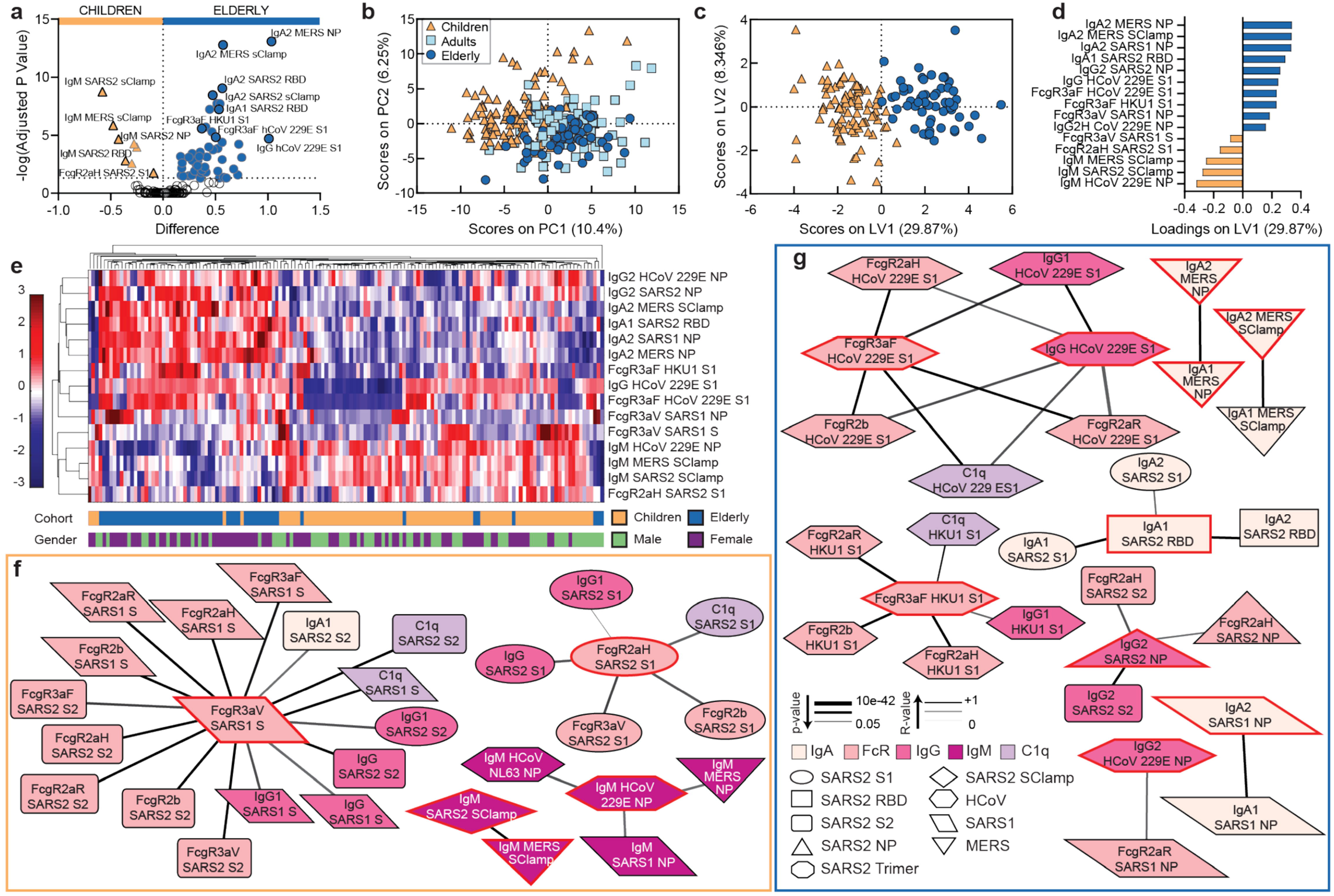
Vastly different serological signatures between children and elderly. (a) Volcano plot of children (orange) versus elderly (dark blue), open circles are not significantly different between two groups. Data was zscored prior to analysis. (b) PCA of all 196 Ab features for healthy children, adults (light blue square), and elderly. PLSDA scores (c) and loadings plots (d) using the 15-feature Elastic-Net selected signature for children versus elderly (0.88% calibration error, 1.44% cross-validation error). Variance explained on each LV is in parentheses. Statistically significant separation of groups was determined using a two-tailed t-test on LV1 scores *p*<0.0001, t = 21.60 (e) Hierarchical clustering of Elastic-Net selected features for children and elderly. Levels are coloured from low (dark blue) to high (dark red). Correlation network analyses for children (f) and elderly (g) identify features associated with the Elastic-Net selected features (red outline). Coded by Ab feature type (colour), antigen (shape), correlation strength (line thickness, alpha <0.05), and correlation coefficient (line colour).

We compared CoV Ab responses between children and elderly, identifying 58 of 196 (29.6%) Ab features as significantly different, taking into account multiple comparisons (all *p*<0.00037; **Figure 1a; Extended Data Table 2**). The volcano plot illustrates the eight Ab features (left) elevated in children, consisting of IgM targeting a range of CoV antigens, including several SARS-CoV-2 antigens (S, NP, and the receptor binding domain (RBD), which is involved in binding to host cells and is a key target of neutralizing Abs). Additionally, we observed elevated SARS-CoV-2 Abs that engaged FcγRIIa-H131 soluble dimers, associated with increased ADCP responses^13^. Conversely, 50 significantly elevated features were observed in the elderly, primarily consisting of IgA and IgG against a range of CoV antigens, along with hCoV-specific Abs that can bind soluble FcγRIIIa dimers, associated with increased ADCC^13,14^.

Using systems serology^15^, we observed vastly different CoV serological signatures between children and elderly **(Figure 1)**. Unsupervised multidimensional visualization (Principal Component Analysis; PCA) of all 196 CoV Ab features clearly distinguished children from elderly, with adults spanning these two cohorts although clustering more closely to the elderly **(Figure 1b)**. To identify the minimal signatures of Ab features that best distinguished children from elderly, we performed feature selection (Elastic-Net) followed by a supervised multidimensional clustering analysis (Partial Least Squares Discriminant Analysis; PLSDA). Fifteen Ab features selected by Elastic-Net could accurately discriminate between children and elderly (99.1% calibration, 98.6% cross-validation accuracy). Significant separation of PLSDA scores occurred across the first Latent Variable (LV1) (*p*<0.0001, t = 21.60, LV1-X axis, which accounted for 29.87% of the signature’s total variance; **Figure 1c**). The loadings plot of LV1 (**Figure 1d**) confirmed that children have elevated IgM to a range of CoV antigens, and elevated SARS-CoV-2 Abs that engaged FcγRIIa-H131 soluble dimers, but also SARS-CoV-1 Abs that engaged FcγRIIIa. In comparison, the elderly had elevated IgA, IgG, and FcγRIIIa binding Abs to CoV antigens. To better visualize how these Ab features could distinguish children from elderly, we performed unsupervised hierarchical clustering and illustrate that the same serological signatures naturally cluster the majority of children from the elderly **(Figure 1e)**. Notably, there were no differences in baseline CoV serological profiles between the sexes **(Figure 1e)**, despite males being clinically associated with higher mortality and more severe COVID-19^4^. Correlates of Ab signatures were further confirmed by feature selection followed by multivariate regression analysis (Partial Least Squared Regression; PLSR), including all healthy individuals to analyse correlates across age **(Extended Data Figure 2)**.

To interrogate Ab functionality and cross-reactivity between antigens of selected CoV signatures, we conducted a correlation network analysis, focusing upon significant correlations of Ab features selected by Elastic-Net. The children’s network **(Figure 1f)** demonstrates how SARS-CoV-2 Abs that engaged FcγRIIa-H131 are associated with SARS-CoV-2 IgG, specifically of IgG1 subclass. SARS-CoV-2 FcγRIIa-H131 immune complex formation was also significantly correlated with multiple other SARS-CoV-2 Fc responses, including FcγRIIb, C1q, and FcγRIIIa. Of interest, SARS-CoV-1 Abs that engaged FcγRIIa-V158, were highly correlated with SARS-CoV-2 Ab responses, potentially due to their high sequence similarity (77%) **(Extended Data Figure 3)**. A separate highly correlated IgM network of multiple CoV NP antigens was also observed. Collectively, these data suggest that children may have elevated SARS-CoV-2 Abs with the capacity to engage a range of Fc effector functions to SARS-CoV-2 S, in addition to elevated IgM responses to CoV antigens in comparison to elderly. The elderly predominantly had hCoV functional Ab responses to S protein, mediated by IgG1 and elevated IgA1 to SARS-CoV-2 RBD, correlative with IgA1 and IgA2 S and IgA2 RBD responses **(Figure 1g)**. Additionally, they had elevated SARS-CoV-2 NP Abs correlating with FcγR signatures to a range of CoV. Amino acid (aa) alignment analyses of NP and S1 proteins showed that there is 91% (NP) and 77% (S1) aa similarity between SARS-CoV-2 and SARS-CoV-1 proteins, while SARS-COV-2 and MERS share 47% (NP) aa similarity, hCoVs NL63 and 229E share 29% and 26% aa similarity, respectively in NP, and hCoV HKU and 229E share 28% and 27% in S1, respectively **(Extended Data Figure 3c)**, supporting the network analyses. Collectively, these data suggest that children have less exposure to CoV antigens but may have enhanced primary humoral immune responses targeted to SARS-CoV-2 compared to elderly.

### HLA-II alleles influence Ab signatures

HLA class II allele information was available for a subset of the healthy individuals (children n=84, adults n=17, elderly n=10, **Figure 2; Extended Data Table 1**). To determine whether HLA-II alleles contributed to differences in Ab predisposition, we conducted Elastic-Net and PLSDA to distinguish Ab responses between the two most frequently observed HLA-DQB1, -DRB1 or -DPB1alleles in our cohort (**Figure 2a, d and g)**. Intriguingly, HLA-DQB1*03:01 and HLA-DQB1 *06:02 were associated with distinct Ab features (**Figure 2b-c**; calibration 86.4%, and 82.6% cross-validation accuracy), as were HLA-DRB1 *07:01 and HLA-DRB1*15:01 (**Figure 2e-f**; calibration 79.1%, and 76.3% cross-validation accuracy), along with HLA-DPB1*04:01 and HLA-DPB 1*02:01 (82.3% calibration and 72.2% cross-validation accuracy; **Figure 2h-i**). These results suggest that HLA-II allelic repertoires of individuals may affect the development of Ab responses after infection or vaccination, possibly contributing to the variable antigen-specific Ab titers and/or signatures observed across individuals.

**Figure 2.**
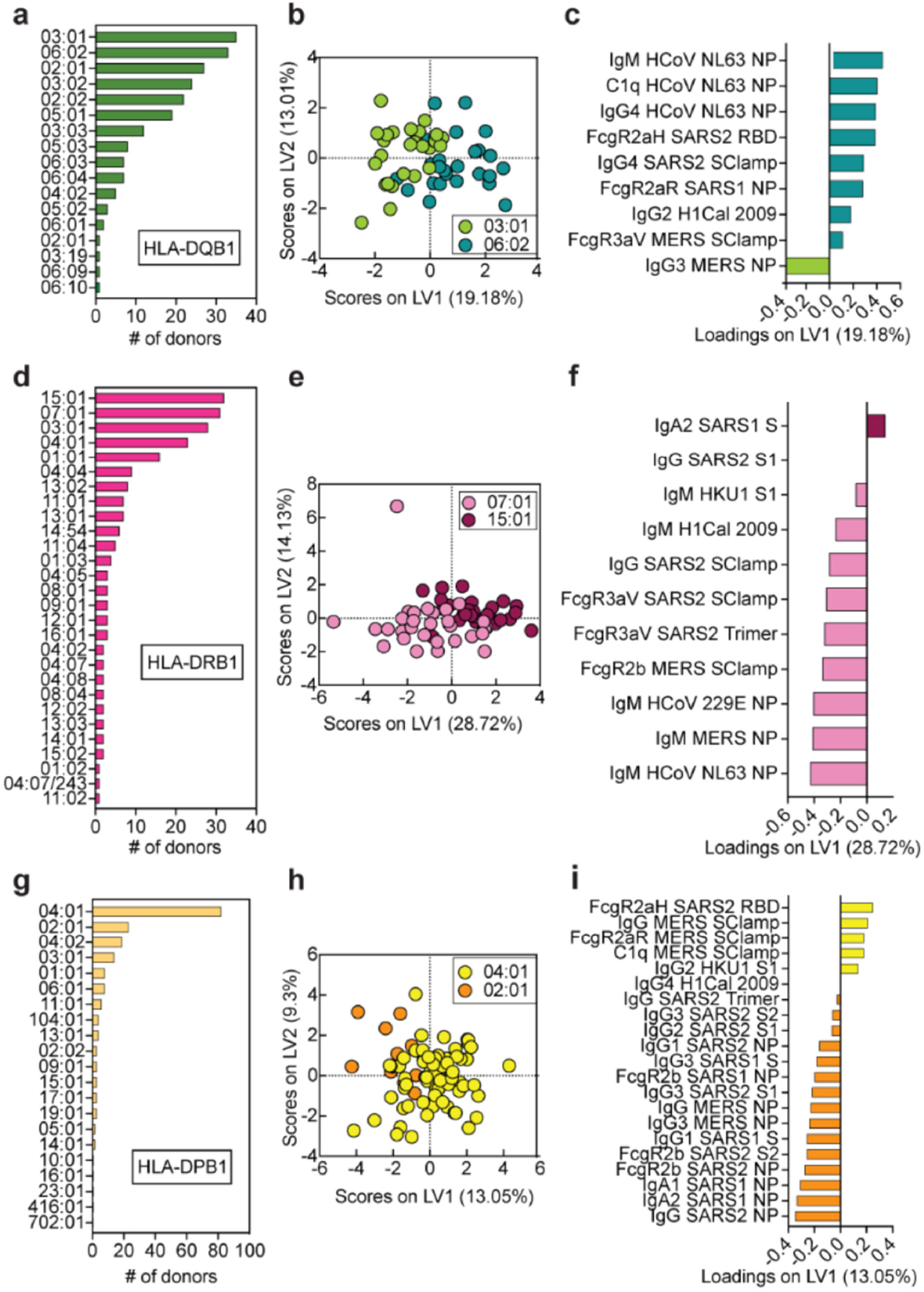
HLA-II alleles influence Ab signatures. HLA-DQB1, -DRB1 and -DPB1 alleles in our healthy donor cohort (a,d,g), PLSDA scores (b,e,h) and loadings (c,f,i) plots using the Elastic-Net selected signatures for the two most frequent alleles (HLA-DQB1*03:01 and 06:02, 17.41% cross-validation error, 13.56% calibration error; HLA-DRB1*15:01 and 07:01, 23.68% cross-validation error, 20.17% calibration error; HLA-DPB1*04:01 and 02:01, 25.78% cross-validation error, 17.66% calibration error). Variance explained on each LV is in parentheses. Analysis was performed on a subset of the healthy individuals (n=111) for whom HLA class II type was available. Donors heterozygotes for the two most frequent HLA alleles were excluded from PLSDA and loading analysis.

### Distinct Fc Ab signature in COVID-19 patients

A cohort of 19 SARS-CoV-2 PCR-positive patients **(Extended Data Figure 2a and Extended Data Table 3)** were screened for SARS-CoV-2 antigen-specific serological profiles **(Extended Data Figure 4)**. An individual who was SARS-CoV-2-exposed but remained SARS-CoV-2 PCR-negative, was also assessed (Donor DD1). Elevated SARS-CoV-2-specific Ab responses in COVID-19 patients relative to healthy or the exposed but PCR-negative individual were observed across multiple titrations **(Extended Data Figure 4)**. In particular, we found that in the majority of COVID-19 patients, the SARS-CoV-2 antigen-specific Abs bound to FcγRIIIaV158 and FcγRIIaH131 soluble dimers at high levels, even at 1:800 plasma titrations, suggesting potent ADCC and ADCP activity.^13,14,16^

Next, we analysed whether COVID-19 patients had distinct serological patterns of SARS-CoV-2 antigen-specific Abs of a single isotype (IgG, IgA, or IgM) compared to healthy individuals (including DD1) using hierarchical clustering. While majority of COVID-19-positive individuals induced high SARS-CoV-2-specific IgM responses, especially to spike antigens, moderate levels of cross-reactive SARS-CoV-2-specific IgM were also detected at high frequencies within the healthy individuals **(Figure 3a)**. Similarly, moderate SARS-CoV-2-specific IgA **(Figure 3b)** and IgG **(Figure 3c)** were observed in healthy individuals, though less frequently than IgM, with cross-reactive IgG responses observed at the lowest frequencies of all isotypes **(Figure 3c)**. Overall, moderate levels of SARS-CoV-2 cross-reactive Abs in healthy donors resulted in poor clustering of COVID-19 patients from healthy individuals when a single isotype were assessed, even though multiple SARS-CoV-2 antigens were included. This suggests that reported low levels of false positives in current serological diagnostics tests could be due to pre-existing levels of cross-reactive Abs that lead to similar serological signatures as observed in SARS-CoV-2-infected individuals when only quantity of antigen-specific Abs are assessed.

To identify the minimum Ab signature that best distinguish the COVID-19 patients from healthy individuals, feature selection was used and identified four SARS-CoV-2 Ab variables that differentiated the two groups **(Figure 3d-f)**, targeting three different SARS-CoV-2 antigens, S trimer, (fold on stabilized spike ectodomain, 2P mutation^17^), NP, and Sclamp (molecular clamp stabilized spike ectodomain^18^). Intriguingly, antigen-specific engagement of FcγRIIIaV158 and C1q, but not IgG, were selected. This suggests that SARS-CoV-2 infection likely induces antigen-specific Ab with distinct Fc qualities, e.g. Fc glycosylation changes, enhancing binding of FcγRIIIaV158 and C1q^19,20^, unlike pre-existing cross-reactive SARS-CoV-2 Abs observed in our healthy donor cohort. In contrast to previous unsupervised hierarchical clustering for IgA, IgM, and IgG **(Figure 3a-c)** to multiple SARS-CoV-2 antigens, these four SARS-CoV-2 Ab features had distinct patterns in COVID-19 patients, which lead to the clustering of COVID-19 patients together with a single exception, this notably being the healthy exposed SARS-CoV-2 PCR-negative individual **(Figure 3d)**. Strikingly, a supervised PLSDA model of these four features, all associated with COVID-19 patients, could significantly distinguish all COVID-19 patients from healthy individuals on LV1 alone (**Figure 3e and f**, *p*<0.0001, t=34.80; 98.51% calibration accuracy, 98.51% cross-validation accuracy). To specifically define these four SARS-CoV-2 Ab features, we conducted a correlation network of Ab responses in the COVID-19-positive individuals **(Figure 3g)**. High levels of correlation were observed between all SARS-CoV-2 spike antigens: S1, S2, RBD, S Trimer, and Sclamp; while NP antigen-specific Ab features created a separate network. Antigen-specific IgG1 and IgG3, which are the most highly functional IgG subclasses^21,22^, were the key mediators of FcγR and C1q antigen-specific Ab engagement. Collectively, these results suggest that future COVID-19 serological diagnostic tests could be improved by assessing the Fc quality of antigen-specific Abs in addition to Ab quantity.

**Figure 3.**
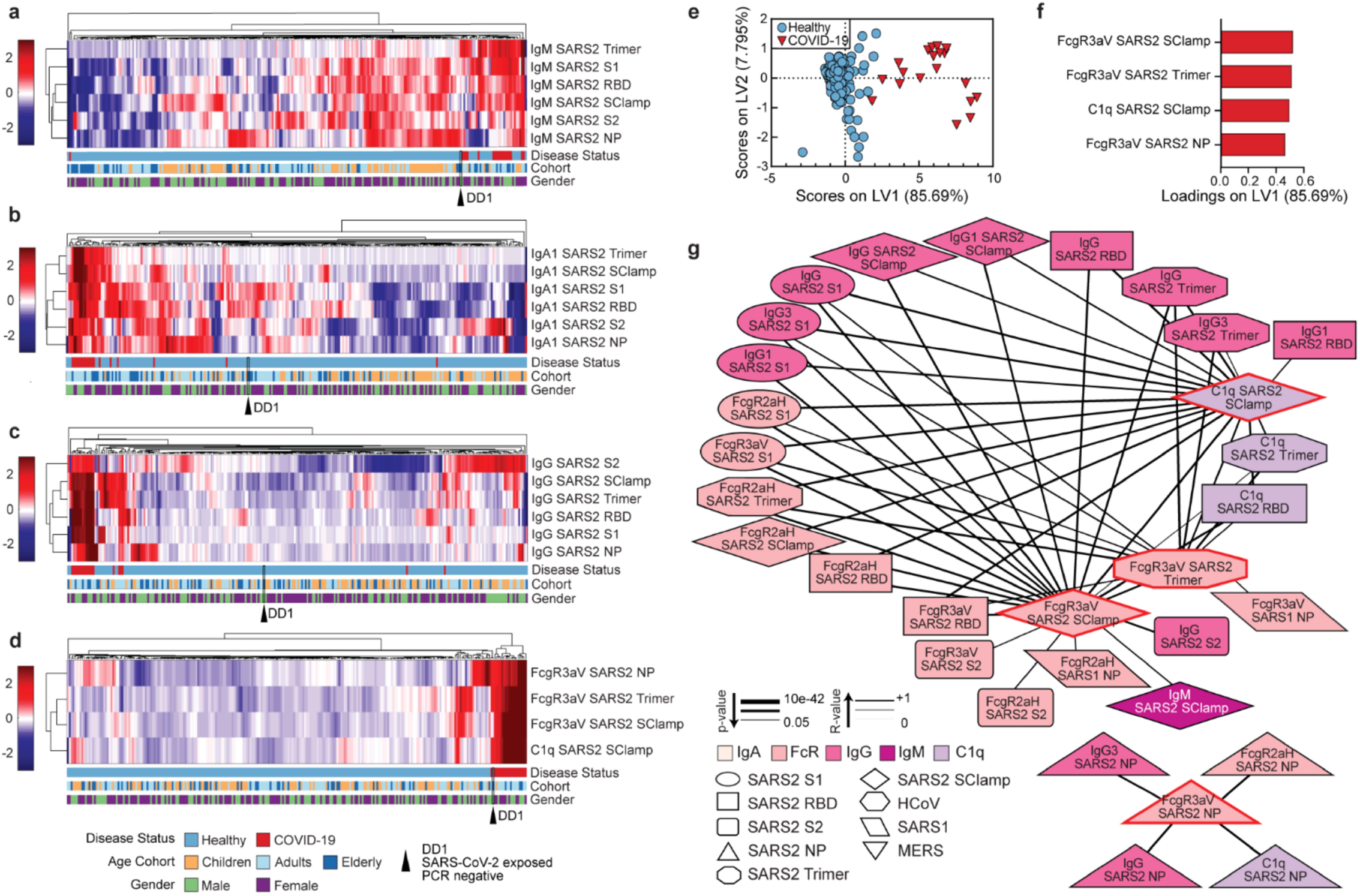
Healthy versus COVID-19 serological signatures. Hierarchical clustering of all SARS-CoV-2 antigens for IgM (a), IgA1 (b) and IgG (c). Levels are coloured from low (dark blue) to high (dark red). Hierarchical clustering (d) and PLSDA model scores (e) and loadings (f) were performed using the four-feature Elastic-Net selected SARS-CoV-2 antigen signature (1.49% calibration error, 1.49% cross-validation error). Variance explained by each LV is in parentheses. (g) Correlation network analysis for COVID-19 patients was performed to identify features significantly associated with the Elastic-Net selected features (red outline). Coded by Ab feature type (colour), antigen (shape), correlation strength (line thickness, alpha <0.05) and correlation coefficient (line colour). Data was zscored prior to analysis.

### Convalescent COVID-19 Ab signatures

The majority of COVID-19 moderate/severe samples were collected upon hospital presentation, whereas mild samples were collected upon convalescence **(Extended Data Table 3)**. Moreover, there was no significant difference between these groups after adjusting for multiple comparisons, which is not surprising given the small sample size **(Extended Data Table 4)**. To explore CoV Ab responses over time, we performed feature selection followed by multivariate regression analysis (PLSR) according to days from first reported disease symptom onset. Not surprisingly, we observed similar Ab signatures to those differentiating healthy from COVID-19 positive individuals, including SARS-CoV-2 antigen-specific Ab engagement with FcγR, C1q, and IgG3 being the most predominant variables associated with convalescence (**Figure 4a-b**, R^2^=0.84, Q^2^=0.72), with several of these Ab features individually associated with days from symptom onset **(Extended Data Figure 5)**. These data are the first indication that SARS-CoV-2 antigen-specific Ab Fc effector functions may have contributed to convalescent of mild/moderate disease.

**Figure 4.**
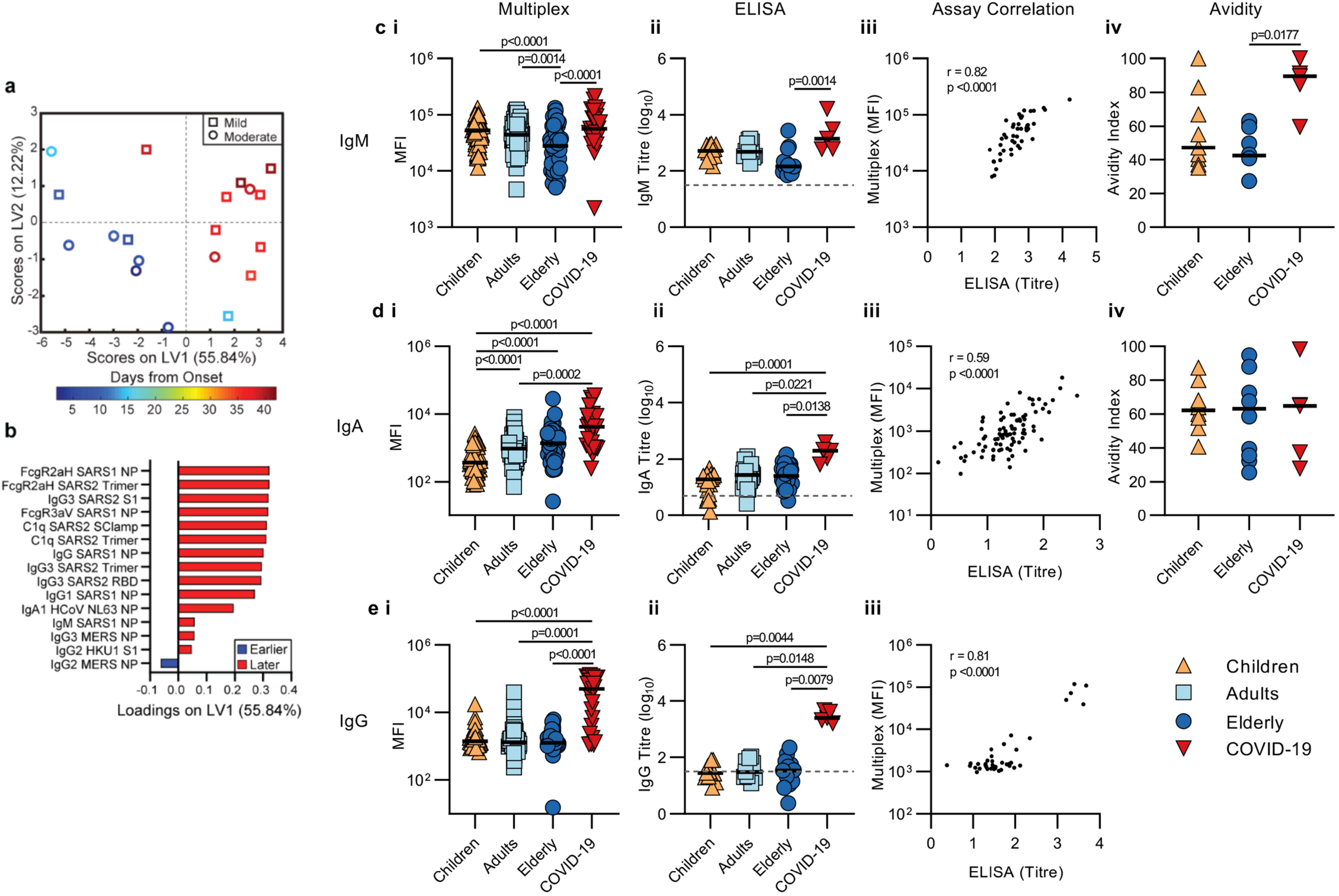
COVID-19 Ab responses over time and RBD Abs in healthy versus COVID-19. PLSR model scores plot (a) loadings plot (b) for all COVID-19 patient data on Elastic-Net 15-feature signature. The model goodness of fit (R^2^) was 0.8361 and goodness of predication (Q^2^) was 0.7194. Percent variance explained by each latent variable in parenthesis. Multiplex MFI data for IgM (c-i), IgA (d-i) and IgG (e-i), ELISA endpoint titers for IgM (c-ii), IgA (d-ii) and IgG (e-ii), and their respective correlations (c-e iii). Avidity index following urea dissociation for IgM (c-iv) and, IgA (d-iv). Children (orange), adults (light blue), elderly (dark blue) and COVID-19 patients (red). Bar indicates the median response of each group. Statistical significance was determined using-Kruskal Wallis with Dunn’s multiple comparisons, exact *p*-values are provided.

### High SARS-CoV-2 RBD-specific IgM in children

Finally, development of neutralizing Ab responses targeting the RBD are associated with control of SARS-CoV-2, occurs in the majority of convalescent COVID-19 serum samples^23^. Therefore, baseline RBD isotype-specific levels between healthy children, adults, elderly and COVID-19 patient plasma samples were assessed via multiplex-assay and validated with published ELISA methods ^24^ **(Figure 4c-i-ii, d-i-ii, e-i-ii)**. Ab detection in both assays was highly correlated **(Figure 4c-iii, d-iii, e-iii)**, confirming earlier observations that children induced elevated IgM, while elderly had higher RBD-specific IgA1 responses as measured by multiplex and trended with ELISA IgA results, while no differences in IgG were observed. Since Ab neutralization quality and potency is often correlated with Ab avidity, we therefore conducted urea disassociation assays on a subset of children, elderly and COVID-19 plasma samples **(Figure 4c-iv, 4d-iv)**. No differences in IgA avidity were found between children, elderly and COVID-19 patients. Avidity of RBD-specfic IgM from elderly was significantly weaker than COVID-19 patients (p=0.0177), while children’s responses, spanning a large range of avidities, were not significantly different (*p=0.0696*). These data, in combination with the overall higher IgM frequency in children, suggest that children may trend to have more potent RBD-specific IgM which may mature more rapidly upon SARS-CoV-2 exposure, as compared to the elderly.

## Discussion

We observed distinct cross-reactive coronavirus serological signatures in healthy children compared to elderly. Children had elevated CoV-specific IgM signatures, whereas elderly had more mature, class-switched CoV-specific IgA and IgG, despite previous epidemiological studies observing higher levels of hCoV infection amongst children compared to the elderly^25^, indicating that multiple rounds of infections over decades might be needed to develop fully experienced CoV humoral immune responses. Intriguingly, school-aged children often have better clinical outcomes during pandemic influenza outbreaks^5^ and can induce more potent broadly-neutralising Ab upon HIV infection^26,27^. It is plausible that upon infection with SARS-CoV-2, the elderly may preferentially induce skewed Ab responses targeting prior cross-reactive hCoV antigens, unlike children who have less experienced immunity and are therefore more likely to mount a more targeted immune response to novel antigens from SARS-COV-2. This immunological phenomenon, known as the Hoskins effect (original antigen sin), observed for several other viral pathogens including influenza and dengue^28,29^ and hypothesized to contribute to enhanced cytokine storm upon dengue reinfection^30^. These hypotheses will need to be confirmed by future studies examining cross-reactive CoV serological signatures from longitudinal sampling of acute SARS-CoV-2-infected children and elderly.

Our results indicate HLA-II alleles contribute to differences in Ab signatures prior to antigen exposure, which could potentially impact antigen-specific Ab profiles upon infection, this field has thus far not been studied and could be associated with antigenic sin. Establishment of an effective humoral immune response after infection and vaccination depends on generation of affinity-matured long-lived plasma cells and memory B cells and is correlated with effective activation of T follicular helper (T_FH_ cells^31^, which depend on the effective presentation of viral epitopes via HLA-II alleles. Several studies demonstrate that variations in HLA-II alleles are associated with susceptibility or resistance to several infectious diseases including MERS-CoV^32,33^ and with vaccine-induced Ab responses^34^. Due to the low frequency of shared HLA-II alleles, this analysis could not be further explored to determine the contribution of age or sex. However, the clear distinction in Ab signatures emphasizes the need to better understand the contributions of HLA-II alleles to the maturation of humoral immunity and would require a sufficiently large cohort of HLA-typed healthy controls and COVID-19 patients.

We also assessed the CoV serological profiles of a small cohort of COVID-19 patients, observing distinct SARS-CoV-2 Fc Ab signatures associated with enhanced engagement of FcγRIIIaV158 and C1q, distinguishing healthy controls from COVID-19 patients. Ab Fc binding to FcγR can be modulated by multiple structural, genetic, and post-translational modifications, including Fc glycosylation^19,35^. Within convalescent plasma samples, we observed upregulation of SARS-CoV-2 antigen-specific IgG, particularly IgG3, which highly correlated with FcγR and C1q engagement, suggesting that Fc functional responses may contribute to recovery. Fc effector functions, while beneficial against many pathogens^15,16,22,36^, can also enhance infection and pathogenesis in other infectious diseases, including dengue, where disease severity is associated with afucosylated IgG1 that enhances FcγRIIIa affinity^37^. This is also observed with other respiratory diseases including tuberculosis, where greater overall inflammation, including inflammatory Fc glycosylation is associated with poorer disease outcomes^36,38^. It is important for future larger SARS-CoV-2 serological studies to assess not only quantitative changes in Ab titers, but also qualitative differences between patients with mild and severe disease, whereas in our study the majority of patients had mild-to-moderate COVID-19.

Overall, our in-depth serological profiling of healthy children, elderly, and COVID-19 patients brings us closer to understanding why the elderly are more susceptible to COVID-19 and provides insights into Ab Fc signatures associated with convalescence of mild/moderate symptomatic individuals. This knowledge is important for the development of improved serological diagnostics, evaluation of convalescent plasma therapeutic trials and will inform immunogenicity assessment of Ab-based SARS-CoV-2 vaccine strategies which could potentially extend beyond neutralizing Abs.

## Data Availability

The source data underlying Figs 1-4, Extended data Figs 1, 2, 4, 5, 6, Extended Data table 2 and 4 are provided as a Source Data file. The coding used for analysis can be found in the Source Coding file. All other data are available from the authors upon request.

## Materials and Methods

### Study participants and sample collection

Our study assessed antibodies to SARS-CoV-2 in a total of 244 healthy individuals and 19 SARS-CoV-2-infected patients **(Extended Data Figure 2a, Table 1 and 3)**. Children undergoing elective tonsillectomy (age 1.5-19) were recruited at the Launceston General Hospital (Tasmania) and, apart from fulfilling the criteria for tonsillectomy, they were considered otherwise healthy, showing no signs of immune compromise. Healthy adult donors (age 22-63) were recruited via the University of Melbourne. Healthy elderly donors (age 65-92) were recruited at the Deepdene Medical Clinic (Victoria). All healthy donors were recruited prior to SARS-CoV-2 pandemic. SARS-CoV-2-infected patients (age 21-75) were recruited at the Alfred Hospital (AH). Convalescent individuals who recovered from COVID-19 were recruited by James Cook University (DD) or University of Melbourne (CP). Eligibility criteria for COVID-19-acute and convalescent recruitment were age ≥18 years old and having at least one swab PCR-positive for SARS-CoV-2. Each patient was categorized into one of the following 6 severity categories: very mild (stay at home minimal symptoms), mild (stay at home with symptoms), moderate (hospitalized, not requiring oxygen), severe/moderate (hospitalized with low flow oxygen), severe (hospitalized with high flow oxygen) or critical (intensive care unit (ICU)). Heparinised blood was centrifuged for 10 min at 300 *g* to collect plasma, which was frozen at −20°C until required. HLA class I and class II molecular genotyping was performed from genomic DNA by the Australian Red Cross Lifeblood (Melbourne).

Human experimental work was conducted according to the Declaration of Helsinki principles and according to the Australian National Health and Medical Research Council Code of Practice. All donors or their legal guardians provided written informed consent. The study was approved by the Human Research Ethics Committee (HREC) of the University of Melbourne (Ethics ID #1443389.4, #2056761, #1647326, #2056689, #1955465) for healthy adult and elderly donors, Tasmanian Health and Medical HREC (H0017479) for healthy child donors, Alfred Hospital (#280/14) for AH donors, James Cook University (#H7886) for DD donors and University of Melbourne (#2056689) for CP donors.

### Deglycosylation of *E. coli*-expressed NP

To minimise background from the *E. coli* expression system, recombinant hCoV 229E and NL63 NP (Prospec-Tany) were first treated with O-glycosidase and PNGase F. Briefly, 40μg of NP were treated with a cocktail of 8μl 10X GlycoBuffer 2, 8μl 10% NP40, 12μl O-Glycosidase, 12μl of Remove-iT PNGase F (New England BioLabs) and water for a final volume of 80μl and incubated at 37°C for two hours on a shaker. The respective mixtures were added to Eppendorf tubes containing 100μl of PSB-washed Chitin magnetic beads (New England BioLabs) to allow the binding and removal of Remove-iT PNGase F. Tubes were agitated for 10 minutes then placed onto a magnetic separation rack for 5 minutes. The supernatant was retrieved and passed through a 100kDa Amicon Ultra centrifugal filter (Merck) to remove remaining O-glycosidase. Finally, NPs were washed with PBS using a 3kDa Amicon Ultra centrifugal filter (Merck) to prepare them for coupling.

### Coupling of carboxylated beads

A custom CoV multiplex assay was designed with SARS-CoV-2, SARS-CoV-1, MERS-CoV and hCoV (229E, HKU1, NL63) S and NP antigens, as well as SARS-CoV-2 RBD (gift from Florian Krammer)^24^, SARS-CoV-2 Trimeric S (gift from Adam Wheatley) and SClamps of both SARS-CoV-2 and MERS-CoV (gift from University of Queensland) **(Extended Data Figure 1b)**. Tetanus toxoid (Sigma) and influenza hemagglutinin (H1Cal2009; Sino Biological), were also added to the assay as positive controls, while BSA blocked beads were included as negative controls. Magnetic carboxylated beads (Bio Rad) were covalently coupled to the antigens using a two-step carbodiimide reaction, in a ratio of 10 million beads to 100μg of antigen, with the exception of the deglycosylated NPs mentioned above in which 40μg were used instead. Briefly, beads were washed and activated in 100 mM monobasic sodium phosphate, pH 6.2, followed by the addition of Sulfo-N-hydroxysulfosuccinimide and 1-Ethyl-3-(3-dimethylaminopropyl) carbodiimide (Thermo Fisher Scientific). After incubation at room temperature (RT) for 30 minutes, the activated microspheres were washed three times and resuspended in 50mM MES pH 5.0 (Thermo Fisher Scientific). The respective antigens were added to the activated beads and the mixture was incubated at RT for three hours on a rotator in the dark. Subsequently, the beads were washed with PBS and blocked with blocking buffer (PBS, 0.1% BSA, 0.02% TWEEN-20, 0.05% Azide, pH 7) for 30 minutes. Finally, beads were washed in PBS 0.05% Sodium Azide and resuspended to one million beads per 100μl.

### Luminex bead-based multiplex assay

The isotypes and subclasses of pathogen-specific antibodies present in the collected plasma were assessed using a multiplex assay as described^39^. Using a black, clear bottom 384-well plate (Greiner Bio-One), 20μl of working bead mixture containing 1000 beads per bead region and 20μl of diluted plasma were added per well. From validation experiments in which cross-reactive Abs present in healthy individuals were titrated, an optimal concentration of 1:100 working dilution of plasma was selected for downstream assays **(Extended data Figure 1c and d)**. The plate was covered and incubated overnight at 4°C on a shaker and was then washed with PBS containing 0.05% Tween20 (PBST). Pathogen-specific antibodies were detected using phycoerythrin (PE)-conjugated mouse anti-human pan-IgG, IgG1-4, IgA1-2 (Southern Biotech), at 1.3μg/ml, 25μl per well. After incubation at RT for two hours on a shaker, the plate was washed, before the beads were resuspended in 50 μl of sheath fluid. The plate was then incubated at RT for 10 minutes on a shaker before being read by the FlexMap 3D. The binding of the PE-detectors was measured to calculate the median fluorescence intensity (MFI). Double background subtraction was conducted, removing first background of blank (buffer only) wells followed by removal of BSA-blocked control bead background signal for each well.

For the detection of IgM, biotinylated mouse anti-human IgM (mAb MT22; MabTech) was added at 1.3μg/ml, 25μl per well. After incubation at RT for two hours on a shaker, the plate was washed, and streptavidin, R-Phycoerythrin conjugate (SAPE; Invitrogen) was added at 1μg/ml, 25μl per well. The plate was then incubated at RT for two hours on a shaker before being washed and read as mentioned above. For the detection of FcγR, soluble recombinant FcγR dimers (higher affinity polymorphisms FcγRIIa-H131, lower affinity polymorphisms FcγRIIa-R131, FcγRIIb, higher affinity polymorphisms FcγRIIIa-V158, lower affinity polymorphisms FcγRIIIa-F158) were provided by Bruce Wines and Mark Hogarth. For the detection of C1q, C1q protein (MP Biomedicals) was first biotinylated (Thermo Fisher Scientific), washed and resuspended in PBS and tertramerized with SAPE. Dimers or tetrameric C1q-PE were added at 1μg/ml, 25μl per well, incubated at RT for two hours on a shaker, then washed. For Dimers, SAPE was added at 1μg/ml, 25μl per well, incubated at RT for two hours on a shaker before being washed and read as mentioned above. Assays were repeated in duplicate. A titration of AH0073 was included in the layout of all multiplex array plates as this patient was known to have IgG, IgM and IgA responses **(Extended Data Figure 6)**. These titrations were used to normalize replicate multiplex array plates.

### Enzyme-linked immunosorbent assay (ELISA)

Detection of RBD-specfic antibodies was performed as described in Stadlbauer et al ^24,40^ with the following modifications; Nunc MaxiSorp flat bottom 96-well plates (Thermo Fisher Scientific) were used for antigen coating, blocking performed with PBS containing 10% BSA and serial dilutions performed with PBST containing 5% BSA. For detection of IgG and IgA, peroxidase-conjugated goat anti-human IgG (Fcγ fragment specific; Jackson ImmunoResearch) or alkaline phosphate-conjugated rat anti-human IgA (mAb MT20; MabTech), was used and developed with TMB (Sigma) substrate for IgG or pNPP (Sigma) for IgA. For IgM, biotinylated mAb MT22 and peroxidase-conjugated streptavidin (Pierce; Thermo Fisher Scientific) was used. Peroxidase reactions were stopped using 1M H3PO4 and plates read at 450nm or 405nm on a Multiskan plate reader (Labsystems). All measurements were normalised using a positive control plasma from a COVID-19 patient (AH0073) run on each plate **(Extended data Figure 6)**. Endpoint titres were determined by interpolation from a sigmodial curve fit (all R-squared values >0.95; GraphPad Prism 8) as the reciprocal dilution of plasma that produced ≥15% absorbance of the positive control. A total of 28 donors from each cohort was randomly selected for IgA analysis, 14 for IgG and 10-14 for IgM. All assays also included 5 same COVID-19 patient samples.

### Antibody avidity assay

Avidity of antibodies in plasma samples was measured using urea as the chaotropic agent and only performed on samples with detectable RBD-specific antibodies (IgA and IgM). Following incubation of plasma at a 1:10 dilution (IgA) or 1:100 dilution (IgM) on RBD-coated plates, 6M of urea was added and incubated for 15 minutes. Bound antibodies were then detected using respective secondary detection reagents described above. The avidity index is expressed as the percentage of remaining antibody bound to antigen following urea treatment compared to the absence of urea.

### Statistical Analysis

Children versus the elderly Volcano plot was conducted using Prism 8. Statistical significance determined using the Holm-Sidak method, with alpha = 0.05 adjusted for 196 tests (Ab features). Each feature was analyzed individually, without assuming a consistent SD. The overall multiplex dataset was analysed for normal distribution using the Shapiro-Wilk test by Prism 8 The data were further analysed by SPSS statistics 26 (IBM Corp.) using the Kruskal-Wallis one-way analysis with a Bonferroni correction to determine the p-values, differences between groups were considered significant at an adjusted p-value of 0.000035 **(Extended data Table 2)**. ELISA data was analyzed using Kruskal-Wallis one-way analysis with Dunn’s multiple comparison using Prism 8. Differences between very mild/mild and moderate/severe/critical patients were analysed using the Mann-Whitney test and differences were considered significant at a *p*-value of 0.05 **(Extended data Table 4)**.

### Data Normalization

For all multivariate analysis Tetanus, H1Cal2009 antigens (positive controls) were removed, with the exception of HLA analysis. Any healthy samples with a missing age, or missing Ab features were removed (n=9). When analysing COVID-19 samples with healthy samples, only the features where data was available for all COVID-19 samples were included. COVID-19 samples lacked entire datasets for IgG4, IgA2, FcgRIIaR131, FcgRIIIaF158, and FcgRIIb, thus these detectors were excluded. When COVID-19 samples were analysed based on the time from disease onset, all visit days were used for each patient. In all other analyses when a patient has two visit days only the second visit was used. Right shifting was performed on each feature (detector-antigen pair) individually if it contained any negative values, by adding the minimum value for that feature back to all samples within that feature. Following this all data was log transformed using the following equation, where x is the right shifted data and y is the right shifted log transformed data: y = log10(x+1). This process transformed the majority of the features to having a normal distribution. In all subsequent multivariate analysis, the data were furthered normalized by mean centring and variance scaling each feature using the zscore function in Matlab. For the HLA analysis, the same data normalization methods were used, except that positive controls were included and all samples with any HLA typing were included. Samples with one copy of each most frequent allele were removed to avoid double classification.

### Feature Selection Using Elastic Net/PLSR and Elastic Net/PLSDA

To determine the minimal set of features (signatures) needed to predict numerical outcomes (age, days from symptom onset) and categorical outcomes (age cohort, COVID-19 infection status, HLA Allele) a three-step process was developed based on^41^. First, the data were randomly sampled without replacement to generate 2000 subsets. The resampled subsets spanned 80% of the original sample size, or sampled all classes at the size of the smallest class for categorical outcomes, which corrected for any potential effects of class size imbalances during regularization. Elastic-Net regularization was then applied to each of the 2000 resampled subsets to reduce and select features most associated with the outcome variables. The Elastic-Net hyperparameter, alpha, was set to have equal weights between the L1 norm and L2 norm associated with the penalty function for least absolute shrinkage and selection (LASSO) and ridge regression, respectively^42^. By using both penalties, Elastic-Net provides sparsity and promotes group selection. The frequency at which each feature was selected across the 2000 iterations was used to determine the signatures by using a sequential step-forward algorithm that iteratively added a single feature into the PLSR (numerical outcome) or PLSDA (categorical outcome) model starting with the feature that had the highest frequency of selection, to the lowest frequency of selection. Model prediction performance was assessed at each step and evaluated by 10-fold cross-validation classification error for categorical outcomes and 10-fold goodness of prediction (Q^2^) for numerical outcomes. The model with the lowest classification error and highest Q^2^ within a 0.01 difference between the minimum classification error or the maximum Q^2^ were selected as the minimum signature. If multiple models fell within this range, the one with the least number of features was selected and if there was a large disparity between calibration and cross-validation error (overfitting), the model with the least disparity and best performance was selected.

### PCA

Principal Component Analysis (PCA), performed in Eigenvectors PLS toolbox in Matlab, is an unsupervised technique that was used to visualize the variance in the samples based on all of the measured features. Every feature is assigned a loading, the linear combinations of these loadings creates a principal component (PC). Loadings and principal components are calculated to describe the maximum amount of variance in the data. Each sample is then scored and plotted using their individual response measurements expressed through the PCs. The percent of variance described by each PC is a measure of the amount of variance in antibody response explained by that respective PC. Separation of groups on the scores plot indicates unsupervised separation of groups based on all features.

### PLSDA

Partial Least Squares Discriminant Analysis (PLSDA), performed in Eigenvectors PLS toolbox in Matlab, was used in conjunction with Elastic-Net, described above, to identify and visualize signatures that distinguish categorical outcomes (age cohort, COVID-19 infection status). This supervised method assigns a loading to each feature within a given signature and identifies the linear combination of loadings (a latent variable) that best separates the categorical groups. A feature with a high loading magnitude indicates greater importance for separating the groups from one another. Each sample is then scored and plotted using their individual response measurements expressed through the latent variables (LVs). The scores and loadings can then be cross referenced to determine which features are loaded in association with which categorical groups (positively loaded features are higher in positively scoring groups etc). All models go through 10-fold cross-validation, where iteratively 10% of the data is left out as the test set, and the rest is used to train the model. Model performance is measured through calibration error (average error in the training set) as well as cross-validation error (average error in the test set), with values near zero being best. All models were orthogonalized to enable clear visualization of results. Statistically significant separation of groups on the PLSDA score plots was determined using a two-tailed t-test on LV1 scores in Prism 8.

### PLSR

Partial Least Squares Regression (PLSR), performed in Eigenvectors PLS toolbox in Matlab, was used in conjunction with Elastic-Net, described above, to identify and visualize signatures that distinguish numerical outcomes (age, days from symptom onset). This supervised method assigns a loading to each feature within a given signature and identifies the linear combination of loadings (a latent variable) that best describes the variance in the numerical outcome. As in PLSDA, a feature with a high loading indicates greater importance for describing the variance in outcome. Each sample is then scored and plotted using their individual response measurements expressed through the latent variables (LVs). The scores and loadings can then be cross referenced to determine which features are loaded in association with which numerical outcomes (positively loaded features are higher in positively scoring samples etc). All models go through 10-fold cross validation, where iteratively 10% of the data is left out as the test set, and the rest is used to train the model. Model performance is measured through R^2^ (average goodness of fit in the training set) as well as Q^2^ (average goodness of prediction in the test set), with values near 1 being best. All models were orthogonalized to enable clear visualization of results.

### Hierarchical Clustering

We visualized separation of numerical (age, days from symptom onset) and categorical (age cohort, COVID-19 infection status) outcomes based on their respective signatures using unsupervised average linkage hierarchical clustering of normalized data; Euclidean distance was used as the distance metric.

### Multiple sequence alignment of CoV Spike and NP

FASTA sequences were obtained from Genpept using the accession numbers as provided by the companies **(Extended data Figure 1b)**. Sequences were cut in Jalview 2.10.5 and subsequently aligned using T-Coffee with the default settings. Residues were coloured to display consensus to the S1 and NP of SAR-CoV-2. Distance matrix for multiple alignment similarities were made in Ugene.

### Software

PCA, PLSDA, and PLSR models were completed using the Eigenvector PLS toolbox in Matlab. Hierarchical Clustering and Correlation Networks were completed using MATLAB 2017b (MathWorks, Natick, MA). PCA, PLSDA, and PLSR scores and loadings plots were plotted in Prism version. Statistical analysis were performed in SPSS. Multiple sequence alignment was done in Jalview 2.10.5 and distance matrix for multiple alignments was done in Ugene 1.16.1 (http://ugene.unipro.ru; Unipro, Novosibirsk, Russia).

## Acknowledgments

We thank all the participants involved in the study, Ebene Haycroft and Brendan Watts for Flexmap3D technical assistance. This work was supported by Jack Ma Foundation to KK, AWC and AW, the Clifford Craig Foundation to KLF and KK, NHMRC Leadership Investigator Grant to KK (1173871), NHMRC Program Grant to KK (1071916), NHMRC Program Grant to DLD (#1132975), NHMRC program grant to SJK (#1149990), Research Grants Council of the Hong Kong Special Administrative Region, China (#T11-712/19-N) to KK. AWC is supported by a NHMRC Career Development Fellowship (#1140509), KK by NHMRC Senior Research Fellowship (1102792), DLD by a NHMRC Principal Research Fellowship (#1137285). SJK by NHMRC Senior Principal Research Fellowship (#1136322). CES has received funding from the European Union’s Horizon 2020 research and innovation program under the Marie Skłodowska-Curie grant agreement No 792532. LH is supported by the Melbourne International Research Scholarship (MIRS) and the Melbourne International Fee Remission Scholarship (MIFRS) from The University of Melbourne. JAJ is supported by an NHMRC Early Career Fellowship (ECF) (APP1123673)

## Author Contributions

KJS, CES, BYC, THON, JC, KLF, ACC, DLD, DCJ, SJK, KBA, KK and AWC formulated ideas, designed the study and experiments; KJS, CES, BYC, THON, LR, LH, MK, CYW, RE, HGK, HXT, JAJ, AKW and AWC performed experiments; FA, FK, KC, NM, DW, PY, BW, PMH and AKW contributed unique reagents; KJS, CES, MML, CYL, SKS, BYC and AWC analysed the experimental data; KJS, CES, MML, CYL, SKS, BYC, KK and AWC wrote the manuscript. All authors reviewed the manuscript.

## Additional information

Supplementary information is available for this paper

Correspondence and requests for materials should be addressed to Katherine Kedzierska and Amy Chung.

## Supplementary information

### Figure Legends

**Extended Data Figure 1. Multiplex Assay setup and optimization**

(a-i-iii) Schematic of bead-based multiplex assay. (b) Overview of antigens included in the assay. (c) Multiplex was validated by measuring a subset of healthy samples both in singleplex and multiplex. Strong correlations suggest that multiplexing did not affect measurement of Ab responses, especially at 1:50 and 1:100 dilutions. (d) Serial dilution of a subset of healthy samples against 14 antigens included in the custom multiplex assay to select dilutions where saturation was not observed. A final dilution of 1: 100 showed not only strong correlation between singleplexed and multiplexed antigens, but was also not saturated for most antigens and was selected for subsequent assays.

**Extended Data Figure 2. Age versus CoV Ab responses**

(a) Overview of the healthy donors per age groups and COVID-19 patients. PLSR model scores plot

(b) loadings plot (c) for all healthy patient data on Elastic-Net 11-feature signature. The model goodness of fit (R^2^) was 0.6421 and goodness of predication (Q^2^) was 0.6144. Percent variance explained by each latent variable in parenthesis. (d) Hierarchical clustering of signature in (b-c). (e) Spearman correlation was performed to associate age with the strength of Ab features against the six SARS-CoV-2 antigens.

**Extended Data Figure 3. Multiple Sequence alignment of NP and S1**

Amino acid sequence alignments of the various NP (a) and S1 (b) used in the assay. Fasta sequences were obtained from Genpept and aligned using T-Coffee with default settings. Amino acids were highlighted to show consensus towards the SARS-CoV-2 proteins, with darker shades of blue being used with increasing consensus. (c) Percentage amino acid sequence alignment for NP and S1 between SARS-CoV-2 and SARS-CoV-1, MERS, hCoV NL63, 229E and HKU1.

**Extended Data Figure 4. SARS2 Ab titrations**

Serial dilutions of COVID-19 plasmas against the six SARS-CoV-2 antigens in the assay. Very mild and mild cases were coloured black, while moderate to severe cases were coloured orange. For comparison, two healthy elderly plasma were included (green). DD1, who was SARS CoV-2-exposed but remained SARS CoV-2 PCR-negative, was also included (purple).

**Extended Data Figure 5. Time from symptom onset and Ab responses**

Antibody features against SARS-CoV-2 antigens which associate with the time of onset of COVID-19, as analysed through Spearman correlation, are displayed. Very mild and mild cases are displayed as black squares while moderate to critical cases are displayed as orange diamonds.

**Extended Data Figure 6: ELISA plasma titrations**

Serial dilutions of plasma from healthy children (n=14) (orange), 12 adults (n=12) (light blue), 14 elderly (n=14) (dark blue) and 5 COVID-19 patients (red) tested in IgM (a), IgA (b) and IgG (c) ELISA. Bold red line represents COVID-19 patient AH0073 who was used as a positive control in all multiples and ELISA plates. Dashed lines represent cut-offs (15% of positive control for IgA and IgG; 30% for IgM) used to interpolate end point titers by non-linear regression analysis.

